# Inclusion of the right ventricular muscle bundle during interventricular septal measurement improves diagnostic accuracy for hypertrophic cardiomyopathy

**DOI:** 10.1101/2025.04.16.25325977

**Authors:** Daniel M Spevack, Pragya Ranjan, Yeraz Khachatoorian, Michael Broker, Chan Woo Kim, Kerry Nevin, Mala Sharma, Divya Malhotra, Srihari Naidu

## Abstract

**Introduction:** Measurement of the interventricular septum (IVS) is a key diagnostic and prognostic parameter in the evaluation of hypertrophic cardiomyopathy (HCM). Right ventricular muscle bundles (RVMB) that parallel the IVS complicate septal measurement on both echocardiography and magnetic resonance imaging. Current guideline statements reference left ventricular wall thickness measurements more than 15 mm as part of the diagnostic criteria for HCM. The medical literature lacks published data on the impact of including RVMB as part of the IVS measurement and its influence on diagnostic accuracy for HCM.

**Methods:** We measured the IVS and RVMB separately on echocardiography in 97 consecutive subjects referred for both echocardiography and magnetic resonance imaging (MRI) as part of the initial evaluation for HCM. Subjects were categorized as having or not having HCM based on current practice guidelines. Patients with HCM were sub-categorized as having septal involvement (HCM-Sep) or primarily apical hypertrophy (HCM-Ap). This was done because subjects with obvious HCM-Ap could be diagnosed with HCM irrespective of IVS thickness.

**Results:** Compared to subjects who did not have HCM, those with HCM-Sep had both increased IVS (15.4 ± 2.7 vs 9.8 ± 1.9 mm, p<0.001) and RVMB thickness (5.2 ± 3.1 vs 1.9 ± 1.9 mm, p<0.001). Within the group of subjects that either had HCM-Sep or did not have HCM, inclusion of the RVMB in the septal measurement increased the sensitivity for HCM from 63% to 100%, whereas specificity decreased from 100% to 87%. RVMB thickness more than 5 mm was seen in 46% of subjects with HCM-Sep but was absent in all subjects with HCM-Ap and those without HCM. The RVMB was visible on long axis imaging in 55% of subjects without HCM, 75% of subjects with HCM-Ap and 85% of subjects with HCM-Sep.

**Conclusions:** Inclusion of the RVMB in the measurement of IVS thickness on echocardiography may improve overall diagnostic accuracy for HCM. In addition, RVMB thickness is increased and is more often visible on parasternal long axis imaging in subjects with HCM, consistent with being part of the HCM pathology. This is particularly true in those with HCM-Sep. These data have implications for standardization of echocardiographic and MRI reporting in HCM.

## Background

Measurement of the interventricular septal (IVS) wall thickness is an important parameter used to help establish the diagnosis of hypertrophic cardiomyopathy (HCM) and to prognosticate the clinical course in patients with this condition.^1-6^ Right ventricular muscle bundles (RVMB) arise from the IVS complicating measurement of wall thickness on both echocardiography and magnetic resonance imaging (MRI), Figure 1. The RVMB that is seen arising from the proximal IVS on echocardiographic parasternal long axis views is part of the crista supraventricularis.^7-12^ The crista supraventricularis separates the RV inflow from the outflow, and serves to direct blood flow. As the crista supraventricularis extends distally toward the apex it typically begins to arc toward the RV free wall and becomes known as the septomarginal trabeculation or moderator band.

**Figure 1.**
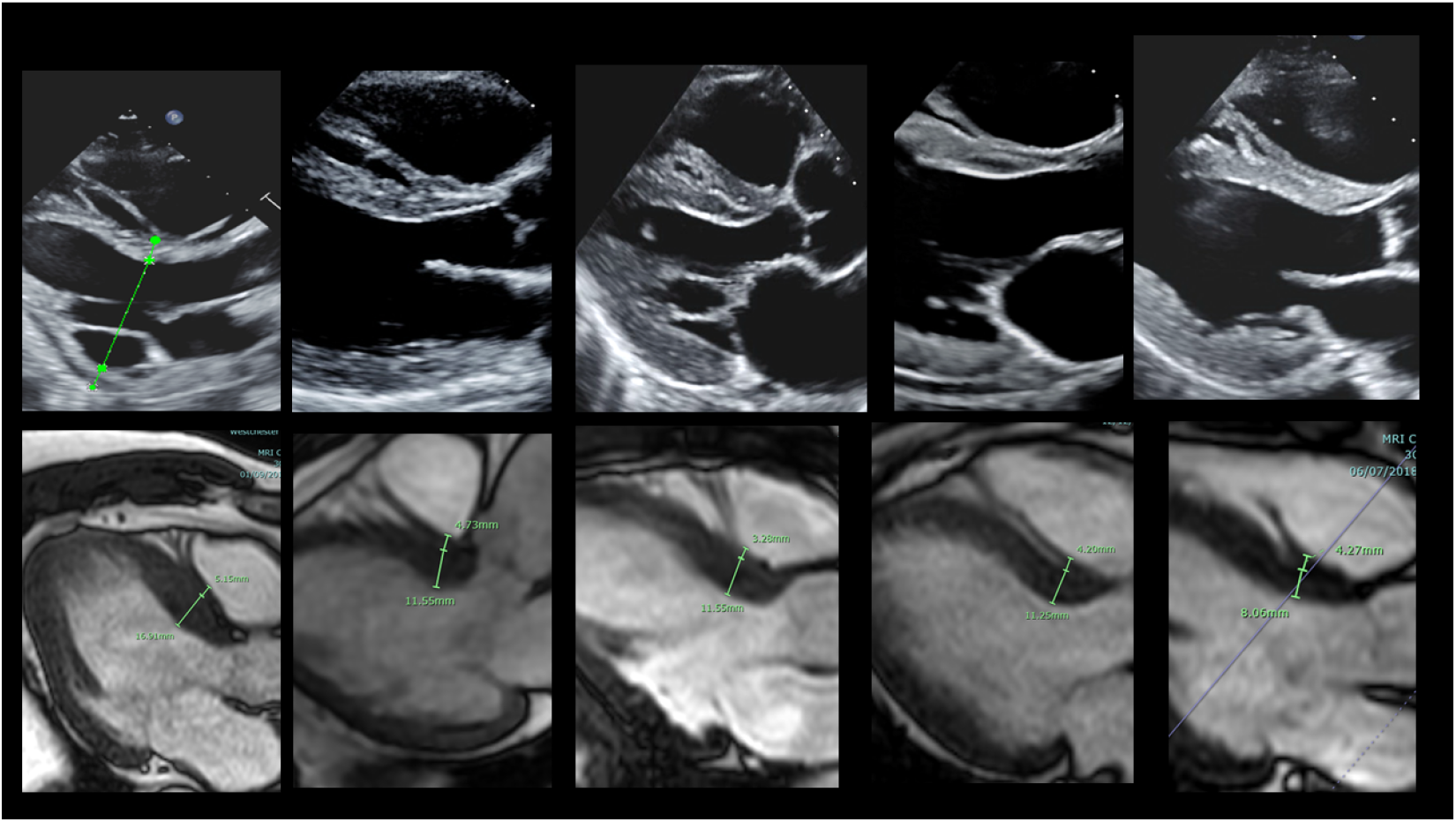
Examples from echocardiography (top row) and magnetic resonance imaging (bottom row) that illustrate the relationship between the primary interventricular septum and the right ventricular muscle bundle (white arrows).

Guideline statements used for both imaging modalities reference increased left ventricular wall thickness more than 15 mm as part of the diagnostic criteria for HCM but are often vague about the need to include or exclude RVMB as part of the of IVS wall thickness measurement.^1-4^ One imaging guideline advises to avoid echocardiographic planes that cut through the wall of the left ventricle tangentially since this could potentially lead to inclusion of trabeculations or papillary muscles in the measurements.^2^ Another statement recommends measuring only compacted myocardium, excluding RV structures.^3^ We are unaware of any prior works that have examined the impact of including RVMB measurement on the diagnostic accuracy for HCM and aimed to examine this practice in a group of subjects undergoing both echocardiography and MRI for evaluation of possible HCM.

## Methods

From our center’s large and nationally certified Hypertrophic Cardiomyopathy Program, the patient roster was queried for all subjects who underwent initial evaluation for HCM that included both echocardiography and MRI over a two-year period. Most MRI had been acquired on Philips Ingenia 1.5 Tesla scanners and were reviewed using IntelliSpace Cardiovascular Information System (Philips Medical Systems, Andover, MA). The MRI reports were reviewed for maximal wall thickness, sites of late gadolinium enhancement and average T1 relaxation times. The electronic health record was reviewed for clinical parameters and medications associated with HCM diagnosis.

Echocardiograms were reviewed for all 97 included subjects. Exams had all been performed on Philips Epiq 7 and Epiq CVx ultrasound machines. Images were viewed using the IntelliSpace Cardiovascular Information System (Philips Medical Systems, Andover, MA). Echocardiographic measurements were performed by a National Board of Echocardiography certified cardiologist (DS) with more than 20 years of experience reading echocardiograms. Measurements of the IVS and RVMB were performed in the parasternal long axis view while blinded to the MRI reports and images. Measurements were made using the reader’s best estimation of the separation between the IVS and RVMB as shown in Figure 2A. If the plane of the parasternal long axis view did not intersect with the RVMB, then the RVMB size was recorded as zero, Figure 2B. While sonographic contrast (perflutren lipid microspheres) was employed in virtually all of the echocardiograms, measurements were made on the non-contrast images unless the septal visualization was suboptimal (n=8). Echocardiograms were also reviewed for systolic anterior motion (SAM) of the mitral valve and left ventricular outflow tract obstruction with and without the patient performing Valsalva.

**Figure 2.**
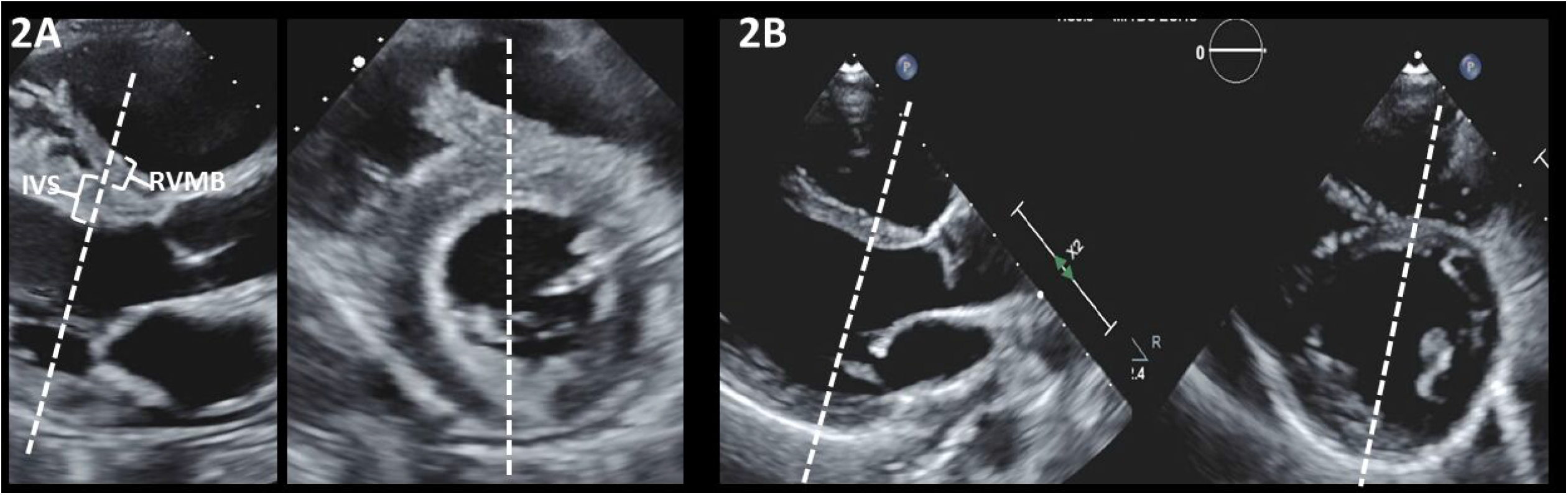
Echocardiographic parasternal long and short axis views. The dashed lines show the plane though which the orthogonal view is likely imaged. In 2A, both views cut through the basal attachment of the right ventricular muscle bundle (RVMB) to the interventricular septum. In 2B, the RVMB can only be seen in the short axis view. The long axis view therefore did not cut through the basal attachment of the RVMB.

Subjects were categorized as having or as not having HCM using the clinical documentation noted by one of the investigators (SN), who is Director of the Westchester Medical Center HCM Program and who had seen each of the study subjects as part of their clinical care. Determination was made using clinical judgement based on noninvasive imaging, invasive assessment, treadmill exercise and clinical criteria in accordance with current AHA/ACC national practice guidelines. Patients with HCM were sub-categorized as having septal involvement (HCM-Sep) or primarily apical hypertrophy (HCM-Ap). This was done because subjects with obvious HCM-Ap could be diagnosed with HCM irrespective of septal wall thickness. The group with HCM-Sep included those with primarily asymmetric septal hypertrophy (n=23), isolated basal septal hypertrophy (n=14) or concentric hypertrophy (n=9).

### Data Analysis

Statistical analysis was done using Stata software, version 11.2 (College Station, TX). Normally distributed data were presented as the mean +/-standard deviation (SD). Non-normal data were presented as the median (range). Comparison of means was performed using the two-sample t-test. Comparison of categorical data was performed using the Chi-squared test. Comparison of medians was performed using the Mann-Whitney test. P-values were considered significant if < 0.05.

## Results

Demographics, clinical characteristics, and imaging findings for the included study subjects are shown in the Table, categorized by pattern of HCM. Compared to those without HCM, subjects with HCM-Sep were older and less often had family history of HCM. On MRI they had higher maximal left ventricular wall thickness and higher global T1 values. On echocardiography, these subjects had increased IVS and RVMB thickness, more often had SAM and had higher left ventricular outflow gradient at rest and with Valsalva. Use of beta blockers, non-dihydropyridine calcium channel blockers, disopyramide and aspirin was also increased in these subjects.

**Table 1.** Demographics, clinical characteristics, and imaging findings for included study subjects categorized by pattern of HCM. P-values are for comparison of group with non-apical HCM to those without HCM.

|  | No HCM<br>(n=31) | HCM-Sep<br>(n=46) | P value | HCM-Ap<br>(n=20) | Total<br>(n=97) |
| --- | --- | --- | --- | --- | --- |
| Age | 43 ± 15 | 54 ± 14 | 0.002 | 56 ± 13 | 50 ± 16 |
| Male sex (%) | 52 | 65 | 0.23 | 65 | 61 |
| BMI | 28 ± 7 | 30 ± 7 | 0.26 | 32 ± 7 | 30 ± 8 |
| Weight (kg) | 85 ± 24 | 91 ± 29 | 0.33 | 94 ± 30 | 90 ± 28 |
| MRI max wt | 11.6 ± 1.9 | 18.8 ± 5.1 | < 0.001 | 16.3 ± 5.2 | 16.0 ± 5.4 |
| T1 global | 1022 ± 35 | 1059 ± 44 | < 0.001 | 1071 ± 39 | 1048 ± 45 |
| Systolic BP (mmHg) | 123 ± 19 | 130 ± 19 | 0.12 | 132 ± 13 | 128 ± 18 |
| Echo bas sept wt (mm) | 9.8 ± 1.9 | 15.4 ± 2.7 | < 0.001 | 12.3 ± 2.5 | 12.9 ± 3.5 |
| Echo RVMB (mm) | 1.9 ± 1.9 | 5.2 ± 3.1 | < 0.001 | 4.0 ± 3.6 | 3.9 ± 3.2 |
| Echo total sept (mm) | 11.7 ± 2.9 | 20 ± 3.9 | < 0.001 | 16.3 ± 5.8 | 16.9 ± 5.7 |
| RVMB > 5mm (%) | 0 | 46 | < 0.001 | 20 | 26 |
| No RVMB seen (%) | 45 | 15 | 0.004 | 25 | 27 |
| SAM (%) | 0 | 54 | < 0.001 | 5 | 27 |
| Resting (mmHg) | 6.4 ± 1.4 | 15.9 ± 15.3 | < 0.001 | 10.6 ± 7.9 | 11.8 ± 11.9 |
| Valsalva (mmHg) | 7.1 ± 3.6 | 30.0 ± 25.3 | < 0.001 | 14.6 ± 14.2 | 19.5 ± 21.4 |
| Hypertension (%) | 34 | 40 | 0.52 | 47 | 37 |
| Syncope (%) | 2 | 7 | 0.22 | 5 | 4 |
| Dyspnea (%) | 4 | 0 | 0.19 | 2 | 2 |
| Afib (%) | 6 | 12 | 0.31 | 0 | 9 |
| Fam Hx HCM (%) | 48 | 21 | 0.008 | 5 | 36 |
| Smoking | 12 | 26 | 0.08 | 16 | 18 |
| Beta blocker (%) | 42 | 79 | < 0.001 | 63 | 60 |
| Ca blocker non- DHP (%) | 2 | 12 | 0.05 | 0 | 6 |
| Ca blocker DHP (%) | 10 | 17 | 0.34 | 21 | 13 |
| ACE/ARB (%) | 22 | 17 | 0.52 | 21 | 19 |
| Diuretic (%) | 16 | 26 | 0.22 | 32 | 21 |
| Aspirin (%) | 20 | 43 | 0.02 | 26 | 31 |
| Statin (%) | 38 | 50 | 0.25 | 58 | 43 |
| Disopyramide (%) | 0 | 12 | 0.01 | 0 | 5 |

Amongst those with HCM-Sep, all had IVS thickness more than 15 mm if the RVMB was included in the measurement on echocardiography. In contrast, when the RVMB was not included in the measurement, 17 of the 46 HCM-Sep subjects had IVS thickness measured less than 15 mm. Amongst those without HCM, four subjects had septal thickness on echocardiography more than 15 mm if the RVMB was included in the measurement. In contrast, none of these 31 subjects had IVS more than 15 mm when the RVMB was not included.

Therefore, within the group of subjects that either had HCM-Sep or did not have HCM, inclusion of the RVMB in the septal measurement increased the sensitivity for HCM from 63% to 100%, whereas specificity decreased from 100% to 87%. Figure 3 shows two examples of subjects with HCM that could potentially be misclassified as not having HCM based on the septal wall thickness if the RVMB was not included in the septal measurement. Figure 4 shows two examples of subjects that did not have HCM that could potentially be misclassified as having HCM if the RVMB was included in the septal measurement.

**Figure 3.**
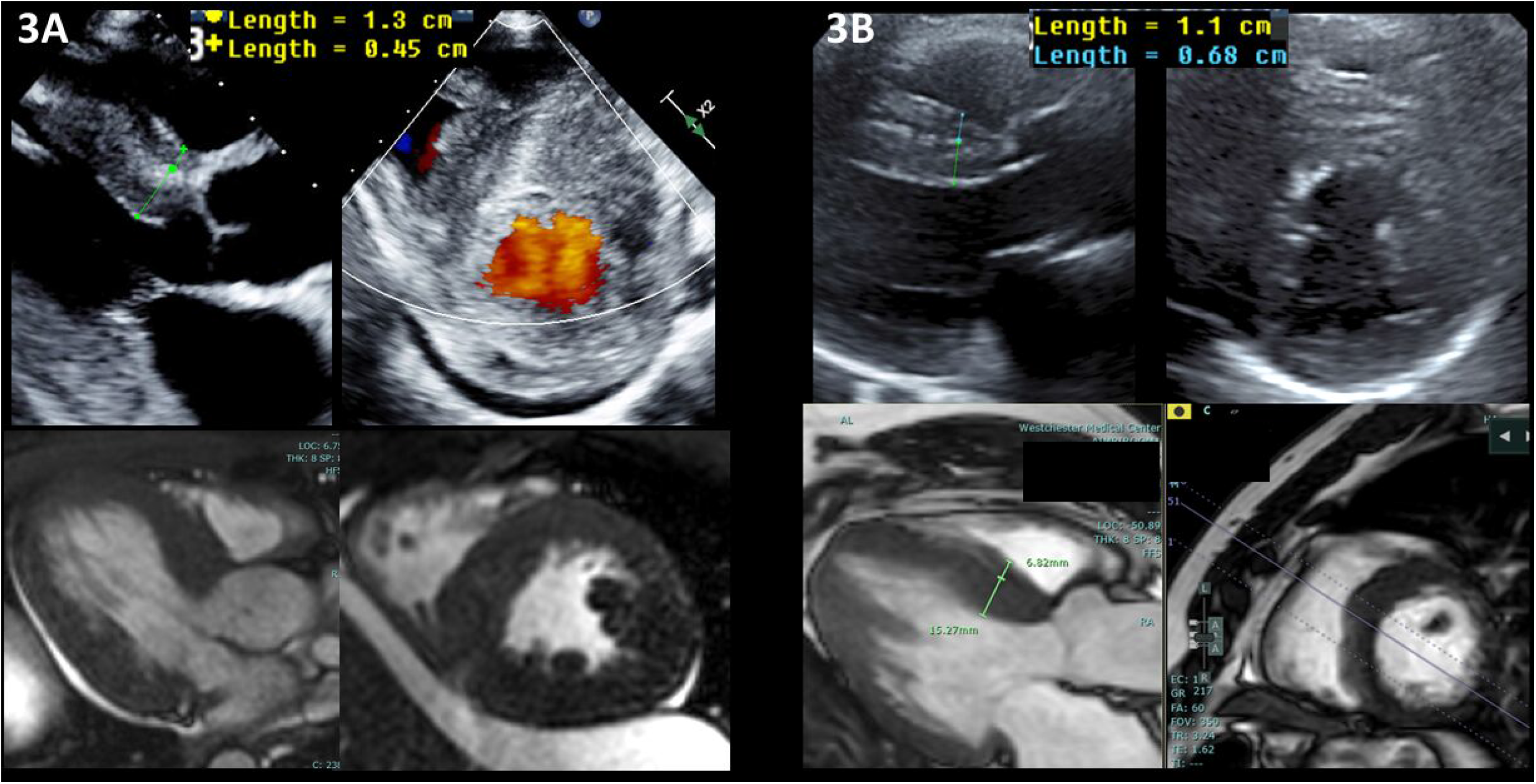
Long and short axis views on both echocardiography (top row) and MRI (bottom row) from two different study subjects (3A and 3B), both with HCM-Sep. In both subjects the primary part of the interventricular septum measured less than 15 mm, but the septal measurement was more than 15 mm when the right ventricular muscle bundle was included.

**Figure 4.**
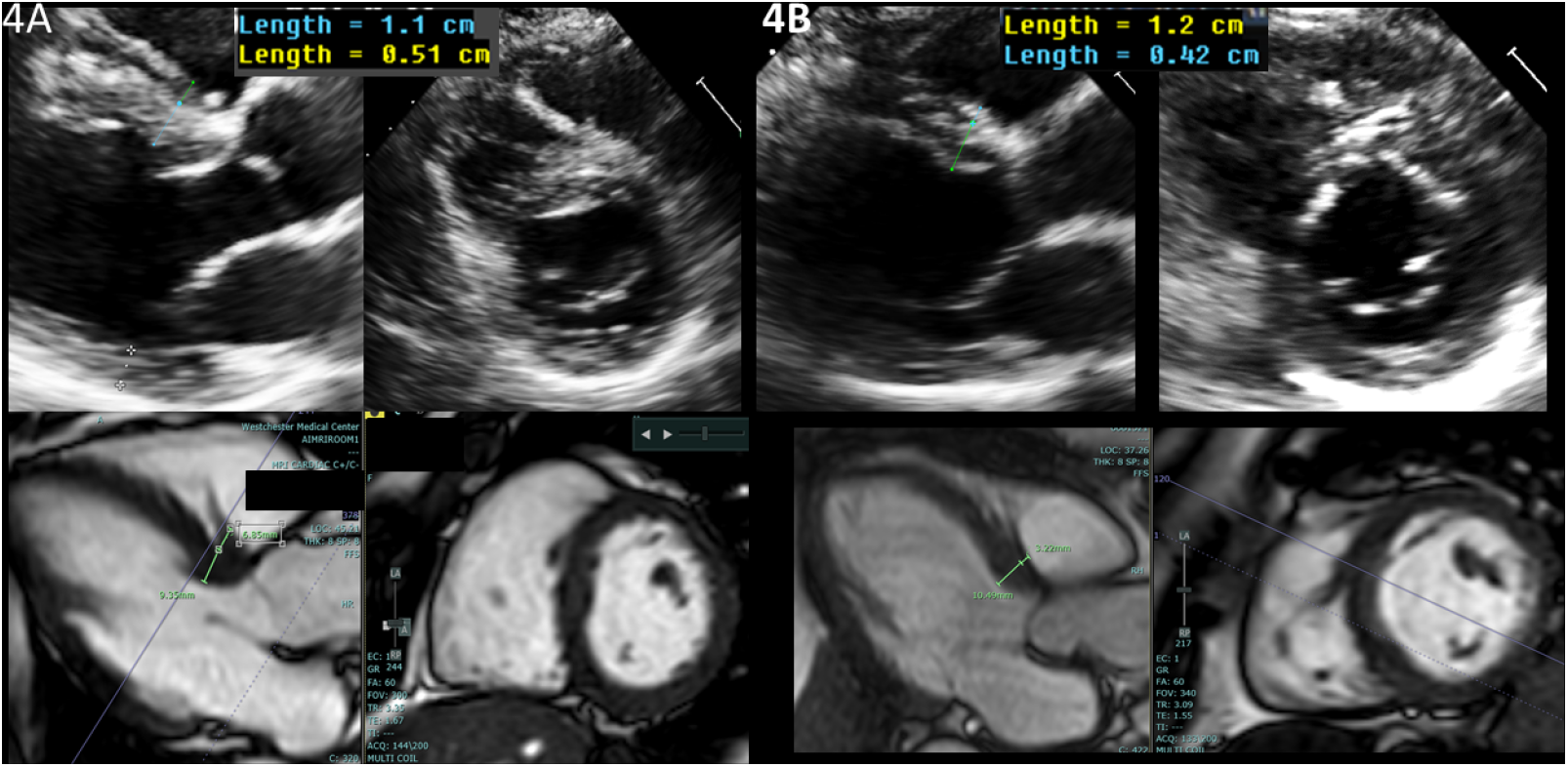
Long and short axis views on both echocardiography (top row) and MRI (bottom row) from two different study subjects (4A and 4B), both without HCM. In both subjects the primary part of the interventricular septum measured less than 15 mm, but the septal measurement was more than 15 mm when the right ventricular muscle bundle was included.

RVMB thickness more than 5 mm was present in 21 of 46 (46%) with HCM-Sep, 4 of 20 (20%) with HCM-Ap and none of the subjects without HCM. Particular interest was paid to subjects in whom primary IVS thickness (ie. without including the RVMB) measured between 11 and 14 mm, since the RVMB measurement had greater impact on HCM diagnosis in this group. For subjects in this group, the RVMB thickness was more than 5 mm in 9 of 16 (56%) with HCM-Sep. In contrast, none of the 12 subjects with HCM-Ap and none of the 16 subjects without HCM had RVMB thickness more than 5 mm.

In some subjects, the plane of the parasternal long axis view did not intersect with the RVMB rendering it nonvisible on that view, Figure 2B. The RVMB was nonvisible in 45% of subjects without HCM, 25% of subjects HCM-Ap and 15% of subjects with HCM-Sep. In the whole study sample, the odds of having HCM were much higher if the RVMB was visible on the echocardiographic parasternal long axis view (odds ratio (OR) 3.7 [95% confidence interval (CI): 1.4, 9.5; p = 0.07]). The odds were even higher if those with HCM-Ap were excluded (OR 4.5 [95% CI: 1.6, 13; p = 0.05]).

## Discussion

The main finding in this study is that subjects with a diagnosis of HCM-Sep had increased thickness of both the primary IVS and the RVMB. Both therefore appear to be pathologically thickened and contributory in HCM. While inclusion of the RVMB in the measurement of the IVS could lead to false positive classification for HCM, in our study sample we found that by including the RVMB the overall diagnostic accuracy was improved. Since echocardiography tends to be the diagnostic imaging tool most commonly employed in the initial evaluation of HCM, it may be wise to use diagnostic criteria with heightened sensitivity by including the RVMB in the IVS wall thickness measurement. MRI can then be employed to confirm the diagnosis in borderline cases at larger referral centers of excellence. We also showed that RVMB thickness more than 5 mm was prevalent in those with HCM-Sep but was absent in those without HCM and in those with HCM-Ap. This may therefore serve as an additional echocardiographic diagnostic criterion for those near the recommended 15 mm cutoff. Another potentially useful finding was that the RVMB was not visible on the parasternal long axis view in a substantial portion of those without HCM, but tended to be visible more often in HCM patients (both HCM-Sep and HCM-Ap). Merely identifying the RVMB in this view greatly increased the odds that the patient may have HCM.

RVMB contribution to IVS thickness measurement could have other important clinical implications beyond the context of HCM diagnostic accuracy. Current HCM guidelines state that IVS wall thickness more than 30 mm is considered a risk factor sudden cardiac death and should lead to consideration of prophylactic implantation of a defibrillator.^1-4^ And since this risk is graded, with lower degrees of IVS thickness (i.e. 25-30 mm) also conferring heightened risk, accurate measurements are helpful for existing risk calculators.^13-15^ We therefore recommend that future investigations address the question of including RVMB thickness during IVS measurement to determine its association with arrhythmia risk. Future study of RVMB and IVS thickness should also be performed in terms of the risk of iatrogenic ventricular septal defect creation following septal reduction therapies with alcohol septal ablation or surgical myectomy.

Our study has several limitations. The patients selected in this study were referred to a large HCM program due to high clinical suspicion for HCM based on prior imaging in another location or a family history of the disease. The diagnostic accuracy estimates for including or excluding the RVMB may be quite different in a sample referred for echocardiography for other indications. Another limitation of this investigation is our categorization of having HCM based on clinical and imaging criteria. Diagnosis of HCM largely based on phenotype is known to have potential for miscategorization. Without routine use of genetic markers and/or tissue diagnosis categorization may have been incorrect in some cases.

In summary, inclusion of the RVMB in the measurement of IVS thickness on echocardiography improves overall diagnostic accuracy, with 100% sensitivity and 87% specificity. RVMB thickness is also markedly increased and more often visible on parasternal long axis imaging in subjects with HCM, both apical and non-apical phenotype. Imaging modalities should be standardized to include measurement of the RVMB in the diagnostic and prognostic evaluation of HCM.

## Data Availability

Anonymized data is available upon request

## References

1. Oomen SR, Mital S, Burke MA, Day SM, Deswal A, et al. 2020 AHA/ACC Guideline for the diagnosis and treatment of patients with hypertrophic cardiomyopathy. Circ. 2020; 142: e558–631.

2. Nagueh SF, Phelan D, Abraham T, Armour A, Desai MY, Dragulescu A, Gilliland Y, Lester SJ, Maldonado Y, Mohiddin S, Nieman K, Sperry BW, Woo A. Recommendations for multimodality cardiovascular imaging of patients with hypertrophic cardiomyopathy: An update from the American Society of Echocardiography, in collaboration with the American Society of Nuclear Cardiology, the Society for Cardiovascular Magnetic Resonance, and the Society of Cardiovascular Computed Tomography. J Am Soc Echocardiogr. 2022; 35:533–69.

3. Mitchell CC, Frye C, Jankowski M, Symanski J, Lester SJ, Gilliland Y, Dragulescu A, Abraham T, Desai M, Martinez MW, Nagueh SF, Phelan D. A Practical approach to echocardiographic imaging in patients with hypertrophic cardiomyopathy. J Am Soc Echocardiogr. 2023; 36:913–32.

4. Maron BJ, Desai MY, Nishimura RA, Rakowski H, Towbin JA, Rowin EJ, Maron MS, Sherrid MV. Diagnosis and Evaluation of Hypertrophic Cardiomyopathy: JACC State-of-the-Art Review. J Am Coll Cardiol. 2022 Feb, 79 (4) 372–389.

5. Maron MS. Clinical utility of cardiovascular magnetic resonance in hypertrophic cardiomyopathy. J Cardiovasc Mag Res. 2012; 14: 1–21.

6. Hindieh W, Weissler-Snir A, Hammer H, et al. Discrepant measurements of maximal left ventricular wall thickness between cardiac magnetic resonance imaging and echocardiography in patients with hypertrophic cardiomyopathy. Circ Cardiovasc Imaging. 2017;

7. Haroun HSW. Comparative anatomy of the cardiac septomarginal trabecula (Moderator band). Anatomy Physiol & Biochem Int J. 2017; 2:

8. Kosinski A, Nowinski J, Dariusz K, Piwko G, Kuta W, Grzybiak M. The crista supraventricularis in the human heart and its role in the morphogenesis of the septomarginal trabecula. Ann of Anatomy. 2007; 447–455.

9. Kosinski A, Kozlowski D, Nowinski J, Lewicka E, Dabrowska-Kugacka A, Raczak G, Grzybiak M. Morphogenic aspects of the septomarginal trabecula in the human heart. Arch Med Sci. 2010; 733–43.

10. Lee JL, Hur M. Morphological classification of the moderator band and its relationship with the anterior papillary muscle. Anatomy & Cell Bio. 2018; 52: 38–42.

11. Wang JMH, Rai R, Carrasco M, Sam-Odusina T, Salandy S, Gielecki J, Zurada A, Loukas M. An anatomical review of the right ventricle. Translational Res Anat. 2019; 17:1–6.

12. Rajiah P, MacNamara J, Chaurvedi A, Ashwath R, Fulton NL, Goerne H. Bands in the heart: Multimodality image review. RadioGraphics. 2018; 39(5).

13. Elliott PM, Poloniecki J, Dickie S, Sharma S, Monserrat L, Varnava A, Mahon NG, McKenna WJ. Sudden death in hypertrophic cardiomyopathy: identification of high risk patients. J Am Coll Cardiol. 2000; 36:2212–2218.

14. Spirito P, Bellone P, Harris KM, Bernabo P, Bruzzi P, Maron BJ. Magnitude of left ventricular hypertrophy and risk of sudden death in hypertrophic cardiomyopathy. N Engl J Me. 2000; 342:1778–1785.

15. Olivotto I, Gistri R, Petrone P, Pedemonte E, Vargiu D, Cecchi F. Maximum left ventricular thickness and risk of sudden death in patients with hypertrophic cardiomyopathy. J Am Coll Cardiol. 2003; 41:315–321.

